# Ten-year mortality among first-time mothers involved in family court care proceedings in England: cohort study using linked, administrative hospital, mortality and family court records

**DOI:** 10.64898/2026.01.15.26344208

**Authors:** Georgina Ireland, Bianca De Stavola, Ruth Gilbert, Matthew A Jay

## Abstract

**Background:** Family court care proceedings are instigated to remove children at risk of harm from parental care. Limited information is available on the health of mothers involved in care proceedings. We assessed maternal mortality and causes of death within ten years of first birth, comparing first time mothers with and without care proceedings.

**Methods:** Using linked, administrative hospital and family court data, we followed a whole-population cohort of first-time mothers delivering between 2007 and 2017 up to ten years. We calculated mortality rates comparing mothers with and without care proceedings. We examined proportions of deaths potentially preventable (suicide, homicide, drugs/alcohol, or injury) and identified factors associated with death after care proceedings.

**Results:** Of 2,775,835 first-time mothers contributing 21,856,503 person-years of observation, 28,405 (1·0%) had proceedings. Following proceedings, 314 (1·1%) died, compared to 5,103 (0·2%) among mothers without (age-standardised mortality ratio: 21·0, 95% CI 14·3, 27·7). Mortality ratios were lowest among first-time mothers aged less than 20 (4·5, 95% CI 3·5, 5·9) and highest for those aged 30-34 (28·3 95% CI 21·3, 37·5). Amongst mothers who died after proceedings, 73% of deaths were potentially preventable compared to 28% among mothers without. Factors associated with death were older maternal age at proceedings, health conditions and court orders related to child removal.

**Discussion:** First-time mothers with care proceedings had 21 times the risk of dying within 10 years than similar-aged mothers. Healthcare, social care and family courts must address the extreme health vulnerability of mothers before, during and after proceedings.

## Background

To prevent significant harm occurring or continuing, child protection services (part of children’s social care in England), universal healthcare services such as maternity, primary care and health visiting, and education services have duties to identify and refer to social care children suspected to be experiencing harm or to be at significant risk of being so due to maltreatment, which includes abuse, domestic violence and neglect. ^1^ Parents in England may lose child custody in such circumstances either through agreement with child protection services or by a mandatory order made by the family court in care proceedings (also known as public law family proceedings). Such court action represents one of the most serious interventions in family life available to the state and results in removal of a child in most instances. ^2^

Maternal mortality is a benchmark for healthcare services and maternal health globally yet longer-term survival is rarely studied. ^3^ Increased mortality among mothers experiencing child custody loss through action by child protective services and/or family court in south London, ^4^ Manitoba^5,6^ and Sweden, ^7^ has been observed, with hazard ratios of death ranging from 2 to 5 comparing those with and without custody loss. Mechanisms that explain this higher risk are likely complex, involving pre-existing poor mental and physical health, ^4–9^ experience of domestic abuse leading to the proceedings as well as a history of abuse and neglect from childhood, the trauma of child removal, ^10^ fear of accessing, or inability to access, public services, including healthcare, ^11^ and lack of support and protection before, during and after proceedings. ^10^

Prioritising resources and planning public health interventions, requires understanding excess mortality among mothers experiencing care proceedings. Previous studies, however, underestimate differences with the general population by using similarly deprived comparison groups (such as those accessing mental health services or siblings without child removal) and controlling for confounders through matching or statistical adjustment. No study has examined all of England. A descriptive study is needed which uses a general population comparison group. ^12^ We therefore aimed to compare ten-year mortality for first-time mothers during the period following care proceedings with similar age mothers who did not experience care proceedings in England.

Our objectives were to: 1) estimate the maternal mortality rate from the birth of the first child according to whether the mother had experienced care proceedings or not; 2) describe the proportions of potentially-preventable deaths (related to suicide, homicide, drug or alcohol use, or accidental injury) and those related to medical or other causes; and 3) identify which factors, including maternal social and health characteristics and proceedings outcome, were associated with mortality after care proceedings.

Findings could inform possible interventions in the early childhood years to improve the health of vulnerable mothers involved in care proceedings. We reasoned that care proceedings indicate health and social vulnerability, regardless of the legal outcome, and that courts, social care (who bring the case to court) and healthcare services have an opportunity and duty to intervene to mitigate risks to the mother.

## Methods

### Population and data sources

We included a cohort of all mothers with a first live birth between 2007 and 2017 and aged 15-39 at the time of birth. We focused on first-time mothers, as early preventive interventions for mothers for their first child can benefit planning for, and outcomes of, subsequent children. The cohort was derived from a broader delivery cohort of all women (15 to 50 years) giving birth in NHS maternity units, available in Hospital Episode Statistics (HES) inpatient hospital records, detailed elsewhere. ^13^ This cohort contained 92·9% of all births registered in England by the Office for National Statistics (ONS) for the 1998 and 2021 period. Mothers were followed for up to ten years following a first live birth recorded in a hospital admission. Hospital admission records contain mandatory diagnostic data using the International Statistical Classification of Diseases and Related Health Problems 10^th^ Revision (ICD-10) for each hospital episode, with one primary diagnosis (the primary cause of the admission) and up to 19 secondary diagnoses. Death registration records include cause of death (ICD-10; underlying cause of death and up to 15 contributory causes).

Administrative data for mothers (15-50 years at start of court case) involved in family court care proceedings in England between April 2007 and March 2022 were linked to the hospital delivery cohort. Data were provided by the Children and Family Court Advisory and Support Service (Cafcass). ^14^ This dataset included, for all family court care proceedings in England, de-identified information on hearing dates, number and ages of children involved in the case, demographic information on the mother, and orders made at the end of the proceedings. Linkage of Cafcass identifiers to HES by NHS England is reported elsewhere (83% of mothers involved in care proceedings in England linked to a mother in the delivery cohort). ^13^

### Outcomes

For objective 1 (mortality rates), the outcome was mortality within ten years of first live birth. Mothers were recorded as having died based on a death recorded in ONS registration records or an inpatient admission whose discharge method or destination was coded as death, with no record of later activity in hospital data. In the case of objective 2 (causes of death), we used cause of death data available in ONS death registration data (underlying cause and up to 15 contributory causes). Cause was not available where death was identified solely on the basis of hospital discharge (0.9% (n=49) of deaths). Additionally, cause of death was missing where there was a pending coroner’s investigation to determine registrable cause for deaths (affecting deaths in 2020 and not completed by 2022, estimated to be fewer than 0.2% of deaths). For objective 3 (factors associated with death following care proceedings), the outcome was mortality within ten years of the start of care proceedings.

### Characteristics

We first described the characteristics of mothers involved and not involved in care proceedings: age at first delivery, racial-ethnic group, Index of Multiple Deprivation quintiles, ^15^ financial year of delivery, presence of health problems in the three years prior to birth. In the whole cohort of first-time mothers (objective 1), and the sub-cohort of those experiencing care proceedings (objective 3), we also categorised health problems in the three years prior to the start of proceedings. Health conditions were identified using previously developed ICD-10 code lists in hospital data. We examined the following overlapping phenotypes: long-term health conditions, ^16^ physical or sensory disability, ^17^ intellectual disability, ^18^ mental health or behavioural conditions, ^4^ and adversity-related admissions (i.e., emergency admissions related to violence, self-harm or drug/alcohol use). ^19^ For objective 3, we classified the final legal order as: (1) case discharge or order of no order; (2) family assessment order or supervision order; (3) residence order, special guardianship order or child arrangements order; (4) care order or secure accommodation order; or (5) placement order. Discharges, orders of no order, family assessment orders and supervision orders usually mean that the child remains at home. In the case of the other orders, the child is likely to have been removed from maternal care.

### Statistical Analysis

For objective 1 (comparing rates of death within ten years of first live birth), we estimated ten-year mortality rates by care proceeding involvement. We followed all mothers from their first live birth until the first of ten years, 51^st^ birthday, 31 December 2020 or death. To address immortal time bias induced by the fact that mothers in the care proceedings group could not have died before experiencing proceedings, person-time before the start of care proceedings was categorised as time not exposed and included in the no-proceedings group.

Crude rates by care proceedings (not—or not yet—experienced *vs* experienced) were calculated as the number of deaths over person-years, overall and in five-year age bands. To account for different age structures in the two groups, we also calculated directly age-standardised mortality rates using the age structure of the whole study cohort as the reference cohort. We calculated age-specific and age-standardised mortality rate ratios. Standardisation was for the whole population of first-time mothers.

For objective 2 (causes of death), we described the proportions of deaths within ten years of first birth that were potentially preventable—i.e., related to suicide, homicide, drugs/alcohol or accidental injury—or related to medical, other or unknown causes, by involvement in care proceedings and age group. Classification was based on ICD-10 codes (available on GitHub, see below). We compared mothers involved and never involved in care proceedings and we used two age groups (<25 or 25+ years at birth) due to small numbers in the finer 5-year age groups.

To address objective 3 (factors at proceedings associated with mortality), the cumulative mortality rate among the care proceedings group was estimated using the Kaplan-Meier method, starting from the start of care proceedings. We report this overall and stratified by age, deprivation at the start of proceedings, health conditions in the three years prior to case start, and final legal order at end of proceedings. We do not present Kaplan-Meier curves due to low cell counts and statistical disclosure control. As our aim was to describe the factors associated with mortality in real world populations, and not elucidate causal mechanisms, we provide unadjusted results. ^12^

Code for creating the deliveries cohort and the cohort used in this study and all data processing and analysis code was written in R 4.4.1. This code and machine-readable copies of the ICD-10 code lists are available on GitHub at https://github.com/UCL-CHIG/deliveries-cohort for creation of the deliveries cohort and https://github.com/UCL-CHIG/cafcass-mortality for further processing and analyses for this study. Small cell counts (<10) and estimates based on these were suppressed as statistical disclosure control.

## Results

We included 2,775,835 women with a first live birth recorded in hospital between April 2007 and December 2017 in England. Of these, 28,405 (1·0%) experienced care proceedings within ten years of their first live birth (median [IQR] 2·9 [1·0, 5·6] years). Mothers involved in care proceedings were younger, more deprived and had more health problems recorded in the 3 years before birth than other mothers (Table 1).

**Table 1.**
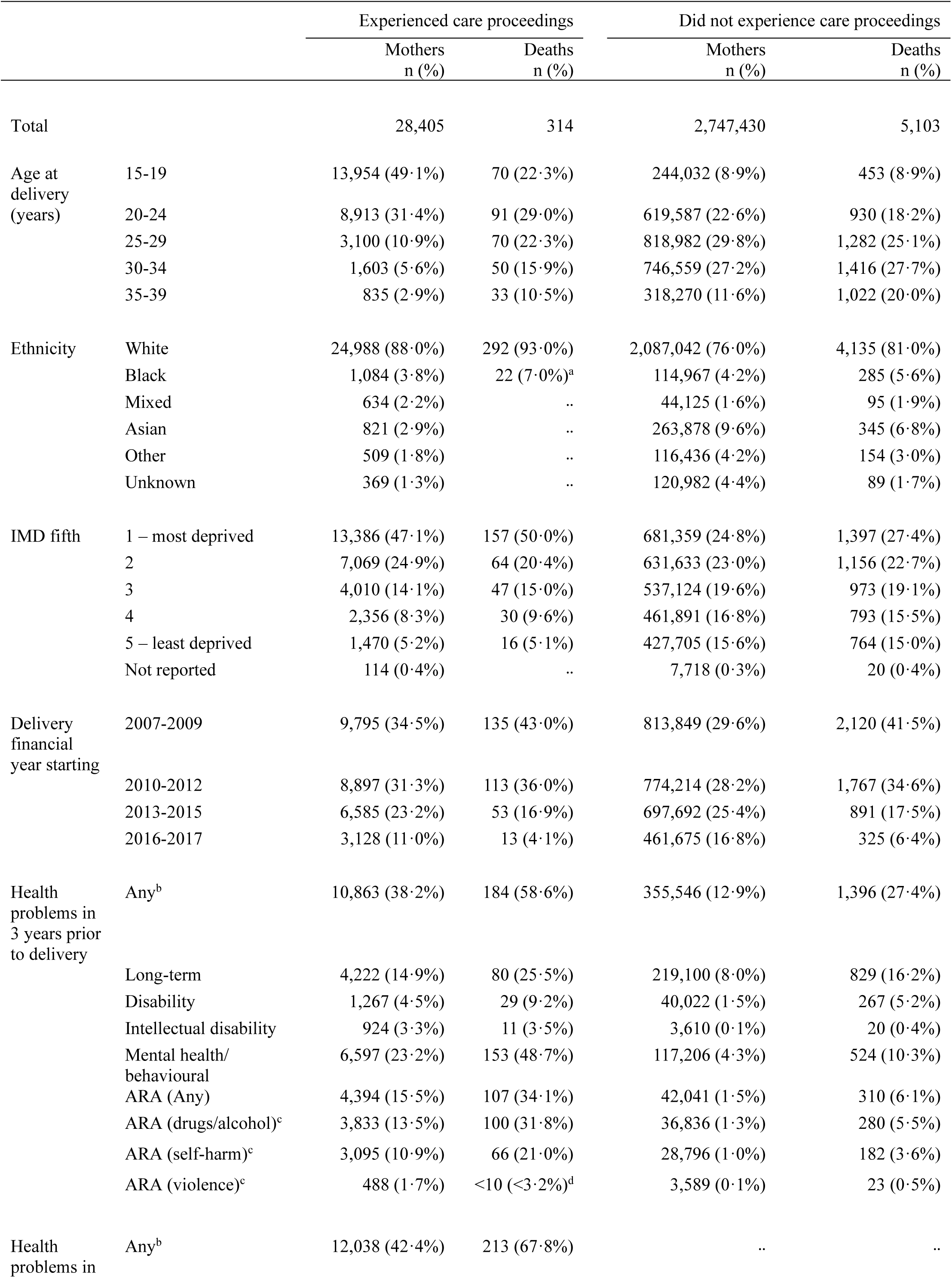
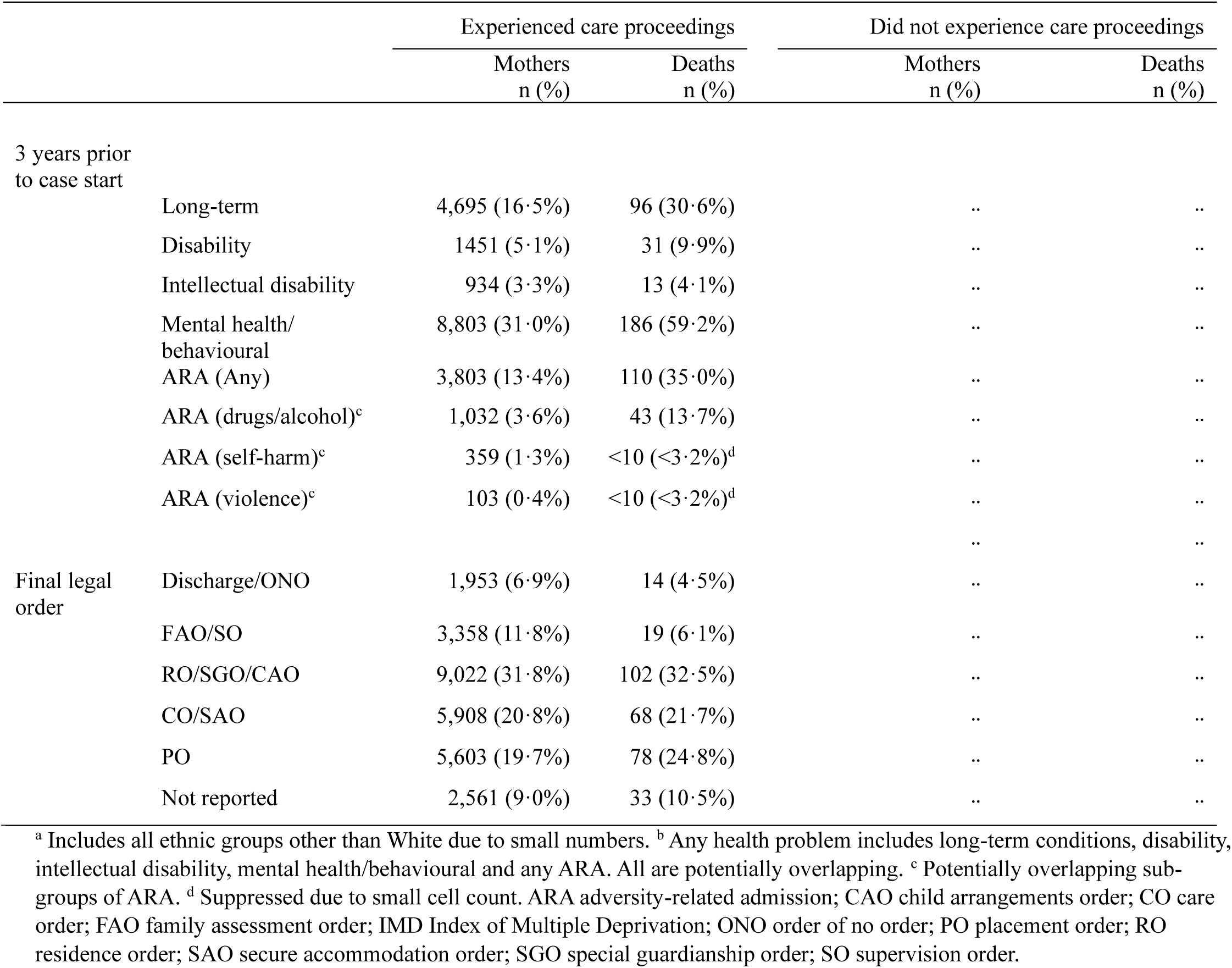
Characteristics of mothers at first birth between April 2007 and December 2017 and number and percent who died within ten years, by involvement in care proceedings or not within ten years.

### Objective 1: mortality from first birth

There were 314 (1·1%) deaths in 28,405 mothers involved in care proceedings and 5,103 (0·2%) in 2,747,430 mothers without care proceedings (median [IQR] follow up time of those in care proceedings 9·5 [6·8, 10·0] years *vs* 8·8 [6·0, 10·0] of those without proceedings) (Table 2).

**Table 2.** Crude mortality rates, crude mortality rate ratios and age-standardised rate ratio for mothers in periods after experiencing care proceedings compared to periods without or yet without care proceedings followed for up to 10 years after first live birth, overall and by maternal age at delivery.

| Periods after care proceedings<sup>a</sup> |  |  |  | Periods without care proceedings<sup>a</sup> |  |  |  |
| --- | --- | --- | --- | --- | --- | --- | --- |
| Age at birth (years) | Person-years | Deaths | Mortality rate (per 1,000 person-years) (95% CI) | Person-years | Deaths | Mortality rate (per 1,000 person-years) (95% CI) | Mortality rate ratio (95% CI) |
| 15-19 | 71,171 | 70 | 0.98 (0.75, 1.21) | 2,093,795 | 453 | 0.22 (0.20, 0.24) | 4.5 (3.5, 5.9) |
| 20-24 | 40,114 | 91 | 2.27 (1.80, 2.73) | 5,021,986 | 930 | 0.19 (0.17, 0.20) | 12.3 (9.9, 15.2) |
| 25-29 | 13,841 | 70 | 5.06 (3.87, 6.24) | 6,429,070 | 1,282 | 0.20 (0.19, 0.21) | 25.4 (19.9, 32.3) |
| 30-34 | 7,254 | 50 | 6.89 (4.98, 8.80) | 5,804,654 | 1,416 | 0.24 (0.23, 0.26) | 28.3 (21.3, 37.5) |
| 35-39 | 4,063 | 33 | 8.12 (5.35, 10.89) | 2,506,998 | 1,022 | 0.41 (0.38, 0.43) | 19.9 (14.1, 28.2) |
| All mothers (crude) | 136,443 | 314 | 2.30 (2.05, 2.56) | 21,856,503 | 5,103 | 0.23 (0.23, 0.24) | 9.9 (8.8, 11.1) |
| All mothers (age-standardised) |  |  | 4.89 (3.37, 6.42) |  |  | 0.23 (0.22, 0.25) | 21.0 (14.3, 27.7) |

<sup>a</sup> To avoid introducing immortal time bias, the follow-up periods before care proceedings are included in the total of person-years without care proceedings. CI confidence interval.

The unadjusted rate of death within ten years of birth for the periods after proceedings was 2·30/1,000 person-years, compared to 0·23/1,000 person-years for the periods without (Table 2). The mortality rate after involvement in care proceedings increased with maternal age at first birth, from 0·98/1,000 person-years in mothers 15-19 years to 8·12/1,000 person-years for mothers 35-39 years (Table 2). By comparison, the mortality rate where mothers had not (or not yet) experienced care proceedings was similar across all ages at around 0·20/1,000 person-years, except for those aged 35-39 years at birth, where it was 0·41/1,000 person-years.

The crude mortality rate ratio comparing periods of time following care proceedings with those without was 9·9 (Table 2). The relationship between increasing age and mortality rate ratio was non-linear, being smallest in the 15-19 age group (4·6) and highest in the 30-34 group (28·3). The age-standardised mortality ratio was 21·0, meaning that if both groups had the same age structure, the rate of death in periods following care proceedings would be 21 times higher than in periods without.

### Objective 2: causes of death

Among mothers with care proceedings, 229 (73%) of deaths were potentially preventable (related to suicide, homicide, drugs/alcohol or accidental injury) compared to 1,405 (28%) among mothers without proceedings (Figure 1; Supplementary Table 1). There was no association with age among the care proceedings group whereas among those without care proceedings, the proportion of potentially preventable deaths was higher among the younger mothers (aged <25 years at first live birth; Figure 1; Supplementary Table 1).

**Figure 1.**
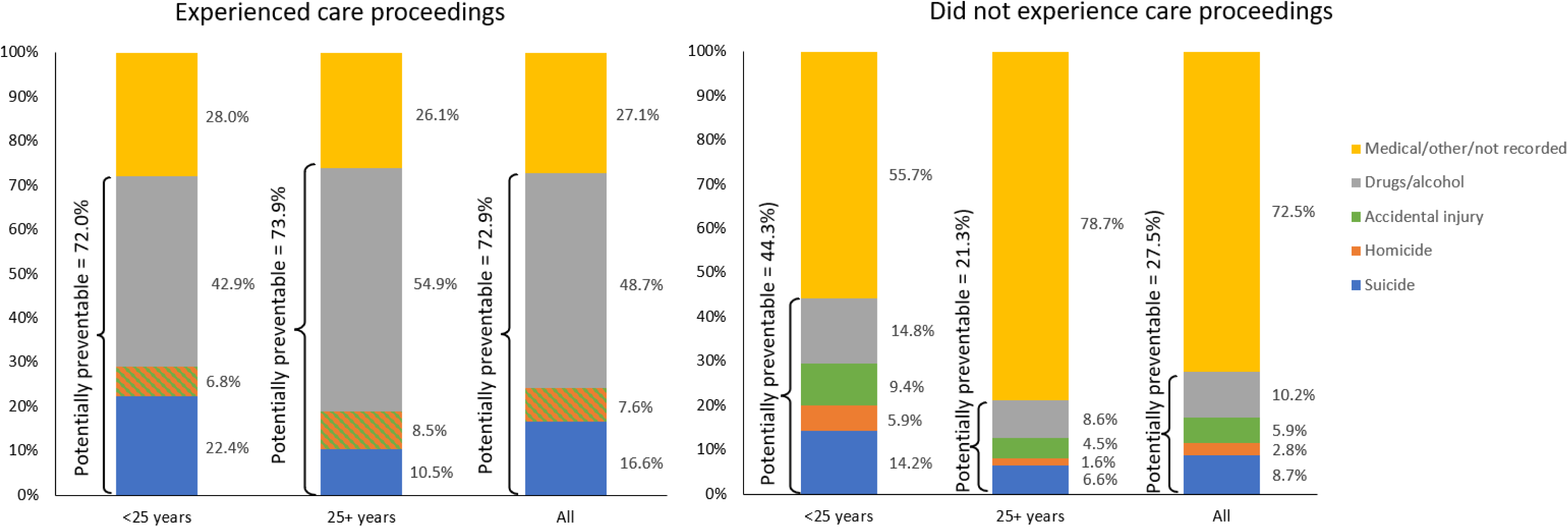
Percentage of deaths related to potentially preventable causes (suicide, homicide, drugs/alcohol, accidental injury) or medical, other or unknown causes by experience of care proceedings and maternal age at first live birth. Deaths due to accidental injury and homicide among mothers who experienced care proceedings are merged due to low cell counts of both. Underlying frequencies are available in Supplementary Table 1.

### Objective 3: mortality following first care proceedings

Kaplan-Meier estimates of cumulative mortality in the ten years following proceedings are given in Figure 2. The overall cumulative incidence was 2·1%. Stratified analyses by various factors highlighted that there were higher cumulative mortality rates with: increasing age at the start of proceedings; pre-delivery health conditions (with the exception of intellectual disability) and adversity-related admissions; and the final legal order at the end of proceedings. In proceedings where the case was discharged, no order was made, or a family assessment order or supervision order was made (i.e., where the child likely remained at home), the cumulative incidence was around 0·8% to 0·9%. This compares to between 2·4% and 2·6% where orders were made implying child removal.

**Figure 2.**
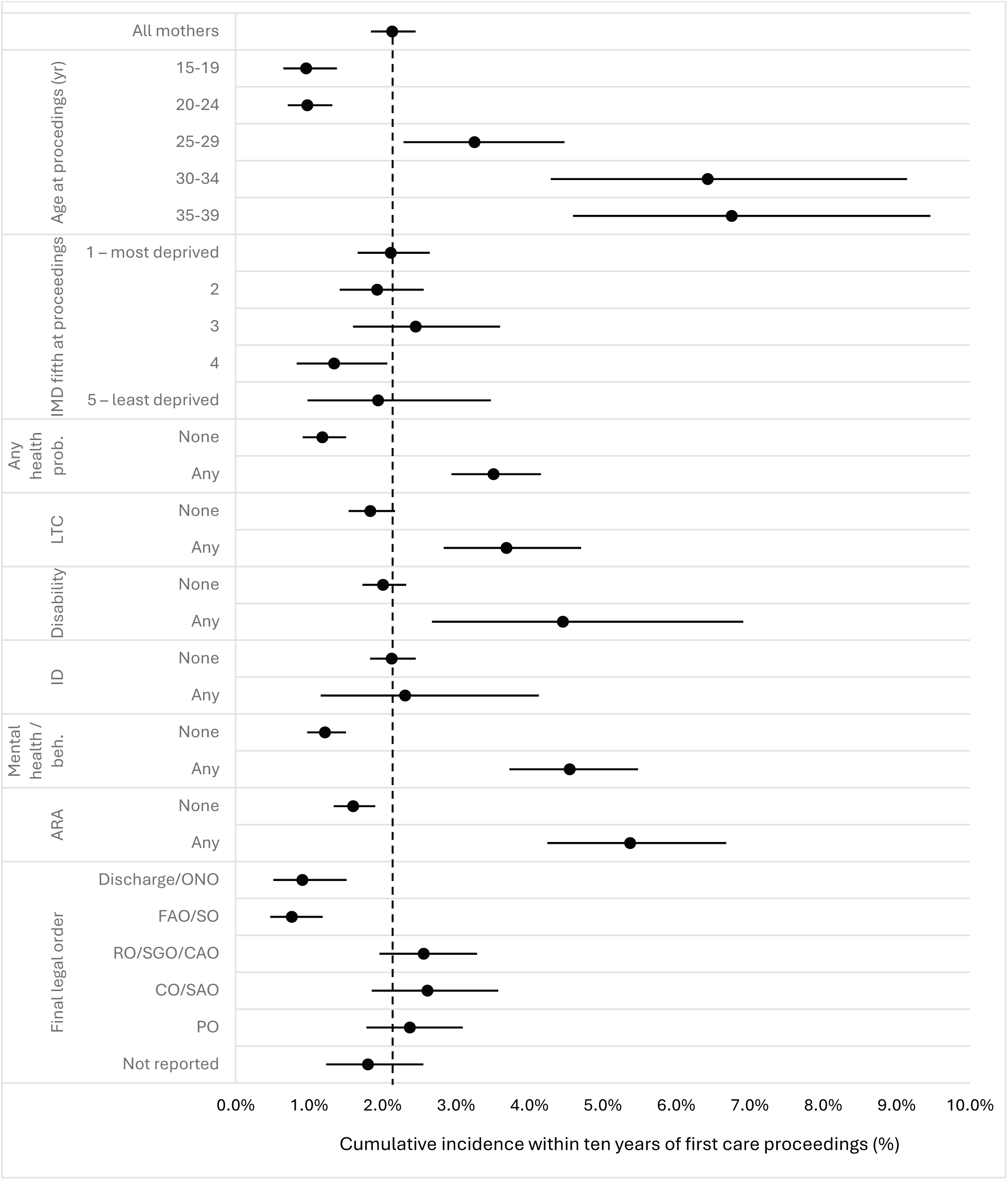
Unadjusted cumulative incidence of mothers’ mortality within ten years of first care proceedings, by age, deprivation, health problems recorded in the three years prior to care proceedings, and final legal order at end of care proceedings. ARA adversity-related admission; beh. behavioural; CAO child arrangements order; CO care order; FAO family assessment order; ID intellectual disability; IMD Index of Multiple Deprivation; LTC long-term condition; ONO order of no order; PO placement order; prob. Problem; RO residence order; SAO secure accommodation order; SGO special guardianship order; SO supervision order. Underlying frequencies are available in Supplementary Table 2.

## Discussion

We found a starkly higher risk of premature death among mothers who experienced care proceedings compared to those who did not. Once age-standardised, and accounting for immortal time bias, women died at a rate 21 times higher in periods following care proceedings than in periods without. Most deaths among the care proceedings group were potentially preventable, being related to suicide, homicide, drugs/alcohol, or accidental injury in 73% of cases, regardless of age. By contrast, among women who had not experienced care proceedings, the percentage of deaths related to these causes was 21% for mothers aged 25 years or older at their first live birth and 44% for mothers aged under 25. The substantially higher percentage of deaths related to these adversity causes and the lack of an association with age further underscore the extreme vulnerability of mothers who experience care proceedings. Our findings suggest that action could be taken to prevent most of these deaths.

As noted in the introduction, mechanisms explaining a higher risk of death are complex, including pre-existing poorer mental and physical health, a theory supported by our data. It may also be that child removal increases maternal mortality risk. ^5–7^ The profound impact of child removal has been described, including long-lasting grief, guilt and stigma; loss of identity as a parent; a sense of failure; and adverse health consequences—all without access to post-removal support. ^10^ Quantitative evidence further indicates outcomes for mothers experiencing custody loss may be worse than those who experience child death. ^6,8,20^ We found that in care proceedings, where orders were made related to child removal, the ten-year mortality risk was between 2·4% and 2·6%, compared to 0·8% to 0·9% where orders were made where the child likely remained at home. The data are therefore consistent with an increased risk of maternal death due to child removal, though such interpretation must be extremely cautious as other contributing factors are likely driving “selection” into these orders, such as more severe previous abuse and health problems. In other words, it is impossible using these data to estimate the causal effect of court decision on maternal mortality. Additionally, no data were available on actual placements (for example, a care order can include a placement at home and between 6% and 9% of children looked after are placed with parents depending on calendar year^21,22^). This could be remedied with linkage to children’s social care data, ^23^ provided these be improved by inclusion of natural identifiers enabling linkage of younger children’s data.

A summary of previous research on mortality risk among similarly deprived mothers can be found in Table 3. ^4–7,24^ Our crude mortality rate (2·3/1,000 person-years) among mothers experiencing care proceedings was slightly lower than the rates of 3·7/1,000 person-years and 3·8/1,000 person-years observed in Manitoba^6^ and Sweden, ^7^ respectively, for mothers with children removed to care (including via non-judicial routes). Our estimated ten-year mortality risk (2·1%) was lower than those of Pearson et al^4^ (among mothers accessing south London mental health services with care proceedings, 3·1%) and Guttmann et al^24^ (among mothers of infants with neonatal abstinence syndrome, 5·1% in England and 4·6% in Ontario). However, the relative difference (age-standardised mortality ratio of 21·0) was higher in our study than in others (hazard ratios of 2·2 to 12·1). In these other studies, comparison groups were similarly disadvantaged, underestimating relative mortality differences vis-à-vis the general population. For example, Pearson et al^4^ compared with other mothers accessing secondary mental health services and Wall-Weiler et al’s^5,6^ comparison groups in Manitoba consisted of biological sisters who did not have children removed or mothers receiving other forms of social care intervention. Additionally, some studies employed matching and adjusted for covariates which likely attenuated differences. We provide unadjusted results, meaning our findings reflect actual real-world risks. ^12^

**Table 3.** Summary of previous studies examining maternal mortality following loss of child custody or in similarly deprived populations.

| Study | Jurisdiction | Cohort and data sources | Exposure group | Comparison group(s) | Findings (rates) | Findings (relative difference) |
| --- | --- | --- | --- | --- | --- | --- |
| Pearson et al (2022) (all-cause mortality: cohort study of women using mental health services) | South London, England | Women using mental health services in four south London boroughs, 2005 to 2020: South London and Maudsley Biomedical Research Centre Case Register linked to Cafcass data | Mothers with care proceedings between 2007 and 2019 and accessing mental health services | Women aged 16-55 years accessing mental health services, matched on type of service accessed and calendar year | Estimated 10-year mortality 3·12% (95% CI 2·47%, 3·94%) among mothers with proceedings and 1·46% (95% CI 1·27%, 1·68%) among women without. | Hazard ratio of death 2·15 (95% CI 1·68, 2·74) accounting for immortal time bias and adjusting for age (mothers with proceedings compared to women without). |
| Guttmann et al (2019) (all-cause and cause-specific mortality: whole population cohort) | England and Ontario, Canada | Whole population cohort of women aged 12 to 49 with a live birth between 2002 and 2012: linked national hospital administrative data and death registrations | Mothers who gave birth to a child with recorded neonatal abstinence syndrome | Mothers who gave birth to a child without recorded neonatal abstinence syndrome | Estimated crude 10-year mortality among exposure group: 5·1% (95% CI 4·7%, 5·6%) in England and 4·6% (95% CI 3·8%, 5·5%) in Ontario. Among controls: 0·42% (95% CI 0·41%, 0·42%) in England and 0·40% (95% CI 0·38%, 0·41%) in Ontario. | Crude hazard ratio for all-cause mortality: 12·1 (95% CI 11·1, 13·2) in England and 11·4 (95% CI 9·7, 13·4) in Ontario; age adjustment did not reduce the hazard ratios. |
| Wall-Wieler et al (2018) (suicide: discordant sibling analysis) | Manitoba, Canada | Women whose first child was born in Manitoba between 1992 and 2015: linked population administrative data including Child and Family Services and mortality data | Mothers with children removed to care by child protective services (which may include non-court-based routes) | (1) a sister with the same biological mother without a child removed to care; (2) mothers without a sister in the cohort who received support from child protective services but did | Suicide attempt rate per 1,000 person-years among those with a child taken into care: 47 (95% CI 37, 49<sup>a</sup>); among sisters: 18 (95% CI 13, 27).<br>Suicide completion rate per 1,000 person-years among those with a child taken into care: 9·5 (95% CI 5·8, 15·6); among sisters: 1·9 (95% CI 0·6, 5·9). | Children taken into care vs sisters (incidence rate ratios): suicide attempt: crude: 2·55 (95% CI 1·68, 3·86); adjusted: 2·15 (95% CI 1·40, 3·30). Suicide completion: crude: 4·99 (95% CI 1·55, 16·02); adjusted: 4·46 (95% CI 1·39, 14·33).<br>Adjusted for age at index date, rurality, suicide attempts in two years prior to index date, number of mood/anxiety disorder diagnoses; number substance use disorder diagnoses. Similar results when comparing mothers with children removed to care with mothers who otherwise received |
|  |  |  |  | not have a child removed |  | support from child protective services but without children removed to care |
| Wall-Wieler et al (2018) (all-cause mortality: discordant sibling analysis) | Manitoba, Canada | Women whose first child was born in Manitoba between 1992 and 2015: linked population administrative data including Child and Family Services and mortality data | Mothers with children removed to care by child protective services (which may include non-court-based routes) | (1) a sister with the same biological mother without a child removed to care; (2) mothers who experienced death of a child | All-cause mortality rate per 1,000 person-years among those with a child taken into care: 3·7 (95% CI 2·9, 4·7); among controls: 1·3 (95% CI 0·9, 2·0). | In fully adjusted models, the hazard ratio of death for all causes comparing women with a child removed to biological siblings without was 3·23 (95% CI 1·62, 6·41). Avoidable mortality: 3·46 (95% CI 1·41, 8·48). Unavoidable mortality: 2·92 (95% CI 1·01, 8·44). Models were adjusted using a family fixed effect and using high-dimensional propensity scores to control for sociodemographic and health factors in the 2 years prior to index date (whether mother had moved, welfare receipt, deprivation, rural/urban, plus over 500 health covariates selected statistically). |
| Wall-Wieler et al (2018) (all-cause mortality: Sweden) | Sweden | Mothers and fathers whose oldest child was born in Sweden between 1990 and 2012: linked population registries | Biological parents who had a child placed into care (which may include non-court-based routes) before 2012 | Biological parents who did not have a child placed into care before 2012, matched on parental year of birth, child's year of birth and child's birth order | Among mothers:<br>All-cause mortality rate per 1,000 person-years among those with a child taken into care: 3·8 (95% CI 3·4 to 4·2); among controls: 0·6 (95% CI 0·6 to 7·0).<br>Among fathers:<br>All-cause mortality rate per 1,000 person-years among those with a child taken into care: 5·7 (95% CI 5·3 to 6·2); among controls: 1·4 (95% CI 1·3 to 1·5). | Among mothers:<br>Adjusted hazard ratios, comparing those with a child taken into care with those without a child taken into care: all-cause mortality: 3·25 (95% CI 2·54, 4·16); avoidable mortality: 3·18 (95% CI 2·36, 4·27)<br>Among fathers:<br>Adjusted hazard ratios (95% CI), comparing those with a child taken into care with those without a child taken into care: all-cause mortality: 1·41 (95% CI 1·20, 1·66); avoidable mortality: 1·59 (95% CI 1·31, 1·94). |

Cafcass Children and Family Court Advisory and Support Service; CI confidence interval. <sup>a</sup> A confidence interval of 38 to 39 is reported, though the upper band is presumably a typographical error.

Strengths of this study include the use of whole population data, providing a cohort of 2·7 million first time mothers all with at least 3 years’ follow-up available for analysis across healthcare, family court and mortality data. Mothers with deliveries in hospital data are representative of all women delivering in England, but the methodology excludes mothers who delivered their first child outside of the NHS, either within private hospitals (<1% of all births), at home (∼2% of all births) or outside of England. Long-term physical or mental health problems are not fully ascertained in hospital data and we likely underestimated the extent to which mothers experienced drug/alcohol use and mental health conditions. To improve case-ascertainment for these conditions, linkages to other datasets are required, including primary care and mental health service data.

Preventive interventions might reduce the need for care proceedings. General practitioners and health visitors are the first point of contact for mothers at their first pregnancy and provide the bedrock of universal healthcare for children and families. These universal healthcare services underpin targeted programmes such as health visiting and the Family Nurse Partnership targeted for first-time teenaged mothers. ^25^ Family support centres, such as Sure Start centres or family hubs, ^26^ may also offer mental health, domestic abuse and substance abuse support, parenting training, or advice about housing and financial problems. However, both universal and targeted services suffer from rising demands, understaffing, and patchy commissioning. For example, a 2022 study reported Family Nurse Partnership commissioning in only two-thirds of local authorities in England, with capacity to enrol only one-quarter of all first-time teenaged mothers intended for support. ^25^ Further, intensive support primarily focuses in the first 1,000 days whereas our findings indicate intensive support may be needed beyond this window and providing support may be difficult where mothers lack trust in public services and/or are stigmatised. ^10^ On-going monitoring and evaluation of mortality and other health outcomes could be supported by routine anonymised linkage between Cafcass and hospital data.

Findings also have implications for family law and policy, including the need to adopt an holistic approach to the family and their problems. The Family Drug and Alcohol Court, which provides a “problem-solving” approach to adjudication in contrast to the family court’s traditional adversarial nature, have shown promise in addressing the underlying problems that led to proceedings in the first place with improved drug use outcomes among parents and higher family reunification rates. ^27^ However, there are only 13 such courts operating in 35 local authorities, which require specialist training and provision. An holistic view should also consider private proceedings (usually between parents following relationship breakdown), which our study did not examine. Those involved in private law proceedings also have poorer mental and physical health and experience deterioration in health as a result of proceedings. ^28^ This is especially so in cases involving domestic abuse, estimated to be the majority of private proceedings. ^28^ Research is needed on mothers involved in both public and/or private proceedings. Such research could also make use of mother-baby links in hospital data^29^ to examine outcomes for children, other datasets including mental health data in England and substance misuse and primary care data in Wales, and data on fathers, who have been understudied to date. Ultimately, a public health approach to family justice is needed that examines why families come to court in the first place, their journeys through the courts and their long-term outcomes following proceedings. ^30^

## Data Availability

Data were shared with the authors by NHS England and Cafcass under data sharing agreement. These data cannot be shared by the study team. Researchers may apply for access to the data for research purposes by contacting NHS England and Cafcass. R code and ICD-10 code lists are available on GitHub at https://github.com/UCL-CHIG/deliveries-cohort and https://github.com/UCL-CHIG/cafcass-mortality.

https://github.com/UCL-CHIG/cafcass-mortality

## Acknowledgements

We are grateful to the Children and Family Court Advisory and Support Service (Cafcass), for providing extracts of their case management data to establish this linkage, and the patients, their families, and NHS staff for their ongoing contribution to research. This work uses data provided by patients and collected by the National Health Service as part of their care and support. Permission to use de-identified data from Hospital Episode Statistics was granted by NHS England (DARS-NIC-196263).

## Authors’ contributions

All authors contributed to conceptualisation and methodology. GI and MAJ contributed to data curation and analysis. MAJ wrote the R code based on initial work by GI. GI and MAJ drafted the manuscript to which all authors contributed in review and editing. GI and MAJ directly accessed and verified the underlying data reported in the manuscript.

## Supplementary Tables

**Supplementary Table 1a.** Number and percentage of deaths by type of death for mothers who experienced care proceedings by age at first live birth (underlying data for Figure 1)

|  | **Experienced care proceedings** |  |  |
| --- | --- | --- | --- |
|  | **<25 years** | **25+ years** | **All** |
| Potentially preventable | 116 (72·0%) | 113 (73·9%) | 229 (72·9%) |
| Suicide | 36 (22·4%) | 16 (10·5%) | 52 (16·6%) |
| Homicide / accidental injury | 11 (6·8) | 13 (8·5%) | 24 (7·6%) |
| Drugs/alcohol | 69 (42·9%) | 84 (54·9%) | 153 (48·7%) |
| Medical/other/not recorded | 45 (28·0%) | 40 (26·1%) | 85 (27·1%) |

Deaths due to accidental injury and homicide among mothers who experienced care proceedings are merged due to low cell counts of both.

**Supplementary Table 1b.**
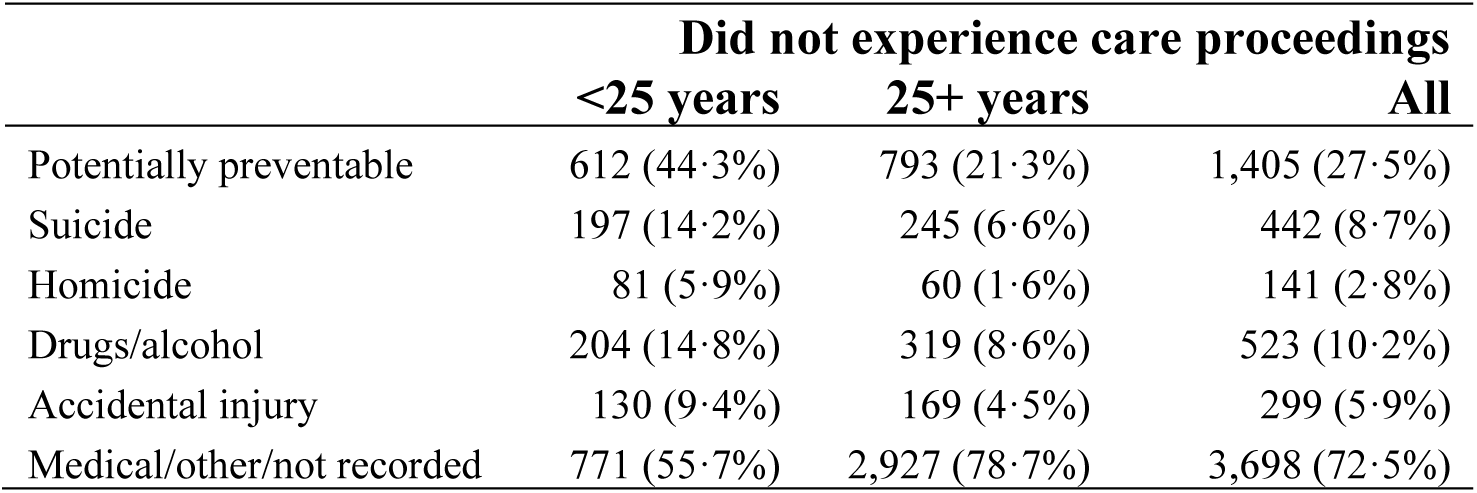
Number and percentage of deaths by type of death for mothers who did not experience care proceedings by age at first live birth (underlying data for Figure 1)

|  | **Did not experience care proceedings** |  |  |
| --- | --- | --- | --- |
|  | **<25 years** | **25+ years** | **All** |
| Potentially preventable | 612 (44·3%) | 793 (21·3%) | 1,405 (27·5%) |
| Suicide | 197 (14·2%) | 245 (6·6%) | 442 (8·7%) |
| Homicide | 81 (5·9%) | 60 (1·6%) | 141 (2·8%) |
| Drugs/alcohol | 204 (14·8%) | 319 (8·6%) | 523 (10·2%) |
| Accidental injury | 130 (9·4%) | 169 (4·5%) | 299 (5·9%) |
| Medical/other/not recorded | 771 (55·7%) | 2,927 (78·7%) | 3,698 (72·5%) |

**Supplementary Table 2.** Number of deaths within ten years of start of care proceedings and unadjusted cumulative incidence of mothers’ mortality within ten years of first care proceedings, by age, deprivation, health problems recorded in the three years prior to care proceedings, and final legal order at end of care proceedings (underlying data for Figure 2)

|  |  | Deaths | 10-year mortality estimate, % (95% confidence interval) |
| --- | --- | --- | --- |
| Total |  | 314 | 2.13 (1.84, 2.45) |
| Age at delivery (years) | 15-19 | 34 | 0.96 (0.65, 1.38) |
|  | 20-24 | 51 | 0.98 (0.71, 1.32) |
|  | 25-29 | 72 | 3.25 (2.29, 4.48) |
|  | 30-34 | 58 | 6.43 (4.29, 9.15) |
|  | 35-39 | 58 | 6.76 (4.60, 9.46) |
| IMD fifth | 1 – most deprived | 129 | 2.11 (1.66, 2.64) |
|  | 2 | 63 | 1.93 (1.42, 2.56) |
|  | 3 | 40 | 2.45 (1.60, 3.60) |
|  | 4 | 21 | 1.34 (0.83, 2.07) |
|  | 5 – least deprived | 12 | 1.94 (0.98, 3.48) |
| Health problems in 3 years prior to case start | None | 101 | 1.18 (0.91, 1.50) |
|  | Any<sup>a</sup> | 213 | 3.51 (2.94, 4.16) |
| Long-term | None | 218 | 1.84 (1.54, 2.17) |
|  | Any | 96 | 3.69 (2.83, 4.70) |
| Disability | None | 283 | 2.01 (1.73, 2.32) |
|  | Any | 31 | 4.46 (2.67, 6.91) |
| Intellectual disability | None | 301 | 2.13 (1.83, 2.45) |
|  | Any | 13 | 2.31 (1.16, 4.13) |
| Mental health/ behavioural | None | 128 | 1.22 (0.97, 1.50) |
|  | Any | 186 | 4.55 (3.73, 5.48) |
| ARA (Any) | None | 204 | 1.60 (1.34, 1.90) |
|  | Any | 110 | 5.37 (4.25, 6.68) |
| Final legal order | Discharge/ONO | 14 | 0.91 (0.51, 1.51) |
|  | FAO/SO | 19 | 0.76 (0.47, 1.19) |
|  | RO/SGO/CAO | 102 | 2.56 (1.96, 3.29) |
|  | CO/SAO | 68 | 2.61 (1.86, 3.58) |
|  | PO | 78 | 2.37 (1.78, 3.09) |
|  | Not reported | 33 | 1.80 (1.23, 2.56) |

<sup>a</sup> Any health problem includes long-term conditions, disability, intellectual disability, mental health/behavioural and any ARA. All are potentially overlapping. ARA adversity-related admission; CAO child arrangements order; CO care order; FAO family assessment order; IMD Index of Multiple Deprivation; ONO order of no order; PO placement order; RO residence order; SAO secure accommodation order; SGO special guardianship order; SO supervision order.

